# N Gene Target Failure (NGTF) for detection of Omicron: a way out for the ‘stealth’ too?

**DOI:** 10.1101/2022.01.28.22269801

**Authors:** Shivangi Harankhedkar, Gaurav Chatterjee, Sweta Rajpal, Afreen Khan, Tuhina Srivastava, Sumeet Mirgh, Anant Gokarn, Sachin Punatar, Nitin Shetty, Amit Joshi, Sudhir Nair, Vedang Murthy, Prashant Tembhare, Ashwini More, Sujeet Kamtalwar, Preeti Chavan, Vivek Bhat, Amar Patil, Sachin Dhumal, Prashant Bhat, Papagudi Subramanian, Rachana Tripathi, Shesheer Munipally, Sumeet Gujral, Navin Khattry, Sudeep Gupta, Nikhil Patkar

## Abstract

S-gene target failure (SGTF) is neither specific nor accurate for identification of Omicron lineage of SARS-CoV-2. We observed N-gene target failure (NGTF) in 402 out of 412 SARS-CoV2 positive cases from December to mid-January 2022 using a commercially available assay. This phenomenon was not observed with more than 15,000 cases tested previously. We sequenced the genome of five samples with NGTF and compared these results with six cases where NGTF was not seen. We confirm that cases with NGTF were the Omicron lineage while cases with preserved N-gene amplification belonged to Delta lineage. We discovered that the ERS31-33 deletion (nucleotide 28362-28370del) overlaps with N gene probe used, explaining NGTF. As the ‘stealth’ Omicron variant also harbors ERS31-33 deletion, this approach will work for the detection of ‘stealth’ Omicron variant as well. We suggest that NGTF can be used as a low cost, rapid screening strategy for detection of Omicron.

## 1. Introduction

The ongoing COVID19 pandemic has witnessed the emergence of novel strains due to the accumulation of mutations in the SARS-CoV-2 genome [1]. Since December 2020, variants of concern (VOC) have been discovered, namely, Alpha (B.1.1.7), Beta (B.1.351), Gamma (P.1), Delta (B.1.617.2). A variant first reported from South Africa was notified to the WHO late in November 2021. Based on genome sequencing this strain was designated with PANGOLIN lineage B.1.1.529 and called the “Omicron” variant (Nextclade strain 21K) [2] [3]. As compared to the original Wuhan strain of SARS-CoV-2, the Omicron variant distinctively harbors a higher number of mutations and is highly transmissible resulting in a higher infectivity rate [4] [5]. This has resulted in a sharp spike in the number of cases all over the globe. This Omicron variant is characterized by more than thirty mutations (including single nucleotide variants as well as insertions and deletions) in the spike protein-coding regions which has raised concerns regarding higher transmissibility and efficacy of vaccines. Furthermore, recent evidence has also emerged that therapeutic monoclonal antibodies have a minimal response against the Omicron variant, presumably due to mutations in the spike protein-encoding regions [5]. This creates the possibility of a clinical need for diagnosis of Omicron variant and distinguishing this from other SARS-CoV-2 variants.

Although the definitive evidence of strain evolution can be obtained by genome sequencing this technique is time-consuming and is available to only a fraction of the laboratories due to the high expertise required to perform this assay. Real-time polymerase chain reaction (RT-PCR) based testing remains the mainstay for relatively rapid testing and community diagnosis.

In India (and presumably all over the world), there is a paucity of commercially available kits that can specifically detect the Omicron variant. It has been suggested that S-gene amplification failure on RT-PCR is a surrogate method to suspect Omicron. However, S-gene amplification failure has also been observed (when using the TaqPath COVID-19 Combo Kit, Thermo Fisher)with the alpha variant of SARS-CoV-2 [6]. In this manuscript, we describe a commercially available RT-PCR assay that can be used as a rapid screening tool to detect the Omicron variant as a result of N-gene amplification failure. We demonstrate sequence level evidence that this approach detects the Omicron variant and reliably distinguishes it from the Delta variant of SARS-CoV-2.

## 2. Methods

Nasopharyngeal and oropharyngeal swabs were received in either viral transport media (VTM) or molecular transport media (MTM) from the fever clinic at the discretion of the treating physician. The testing cohort included hospital staff as well as cancer patients. Real-time polymerase chain reaction (RT-PCR) was performed using a commercially available SARS-CoV-2 assay (Huwel Life Sciences). A total of 15144 tests were carried out over a period of around 23 months (March 2020-mid Jan 2022). Coinciding with the third wave of the pandemic in India, a rise in positivity rate was observed (412 positives out of 1484 samples tested between December 01st 2021 and January 16^th^, 2022, positivity rate of 27.76%) (Figure 1). Eleven cases diagnosed as positive for SARS-CoV-2 by RT-PCR in December 2021 were selectively chosen for viral genome sequencing (five with N-gene amplification failure and six cases without N-gene amplification failure). Amplicon based viral whole-genome sequencing was carried out as per protocols provided by the ARTIC network initiative [7]. Library preparation was performed using the QIAseq FX library kit (Qiagen, Germany) followed by paired-end sequencing on an Illumina MiSeq platform at a targeted depth of 0.3 million reads per sample. For analysis of genomic data, the raw fastq reads were processed for quality checks and parsed through the bioinformatics pipeline for the generation of consensus fasta sequences. Alignment to the reference genome sequence (GenBank: MN908947.3, RefSeq: NC_045512.2; Wuhan-Hu-1 genome) was done using using BWA-MEM algorithm (v.0.7.17). SAM files were processed further using Samtools (v.1.9) and variants were called using iVar (v.1.2) at a minimum alternative frequency of 0.01.

**Figure 1:**
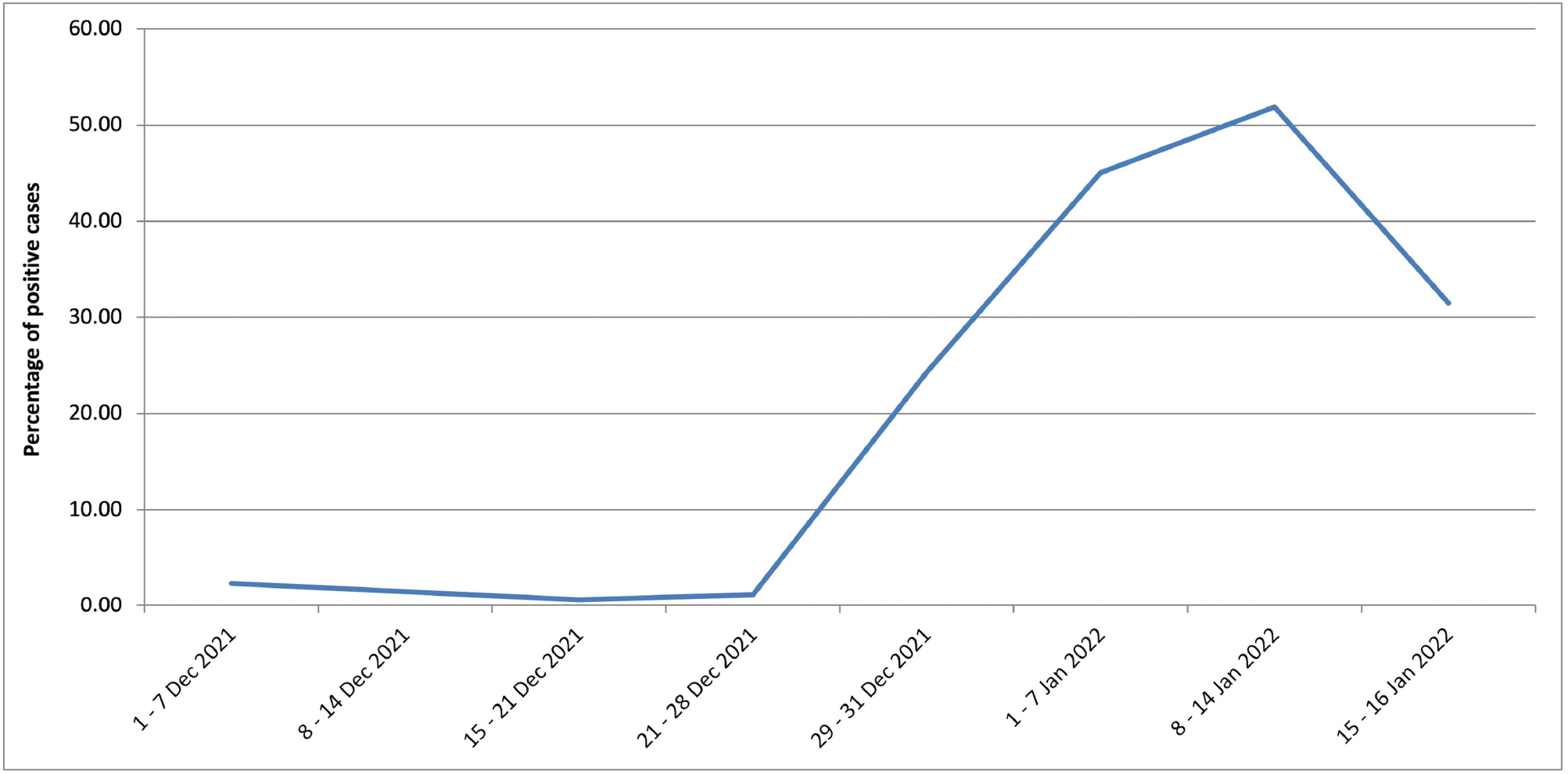
Week wise SARS-CoV-2 positivity rate showing sharp rise in cases towards late December 2021.

Lastly, Nextclade web application was used to process the fasta sequences for identification of clade and mutation calling. The Phylogenetic Assignment of the named Global Outbreak LINeages (PANGOLIN) method of lineage classification was followed [8]. Due approval by Institutional Ethics Committee of Tata Memorial Centre was obtained for this study.

## 3. Results

We have performed over 15000 tests for detection of SARS-CoV-2 RNA, using a commercially available RT-PCR kit (Huwel Life Sciences, Hyderabad, India), which involves the detection of E (Envelope) and N (Nucleocapsid) genes of the virus (Figure 2). A positive result has conventionally required amplification in the E and N gene both. Overall positivity rate of 16.2% was observed during last two years. Towards end of December 2021, as Omicron variant began rising worldwide, we also noticed a sharp rise in the number of positive cases. Positivity rate in January 2022 (1^st^ to 16^th^ January 2022) escalated to 47.24% (386 out of the total 817 samples) from the previous 3.89% in December 2021 (26 positives out of 667 samples, Figure 1). We observed that cases diagnosed from December onwards, showed amplification of E gene alone (N gene target failure, NGTF), a clear divergence from our trend of the past two years (Figure 2). NGTF was observed in 402 out of the 412 positive cases (between 1^st^ of December 2021 to 16^th^ of January 2022), while the remaining ten showed amplification in both E and N genes (Figure 3). On examination of N-gene primer and probe sequences we confirmed that the ERS31-33 deletion (nucleotide 28362-28370del), characteristic of Omicron variant, overlapped with the N gene probe used, resulting in NTGF. We further selected five cases with NTGF, and six cases showing amplification in both E and N genes for genome sequencing. All five cases with NTGF were confirmed on genome sequencing as consistent with Omicron variant (BA.1 lineage), whereas all six cases with both N and E gene amplification belonged to Delta.

**Figure 2:**
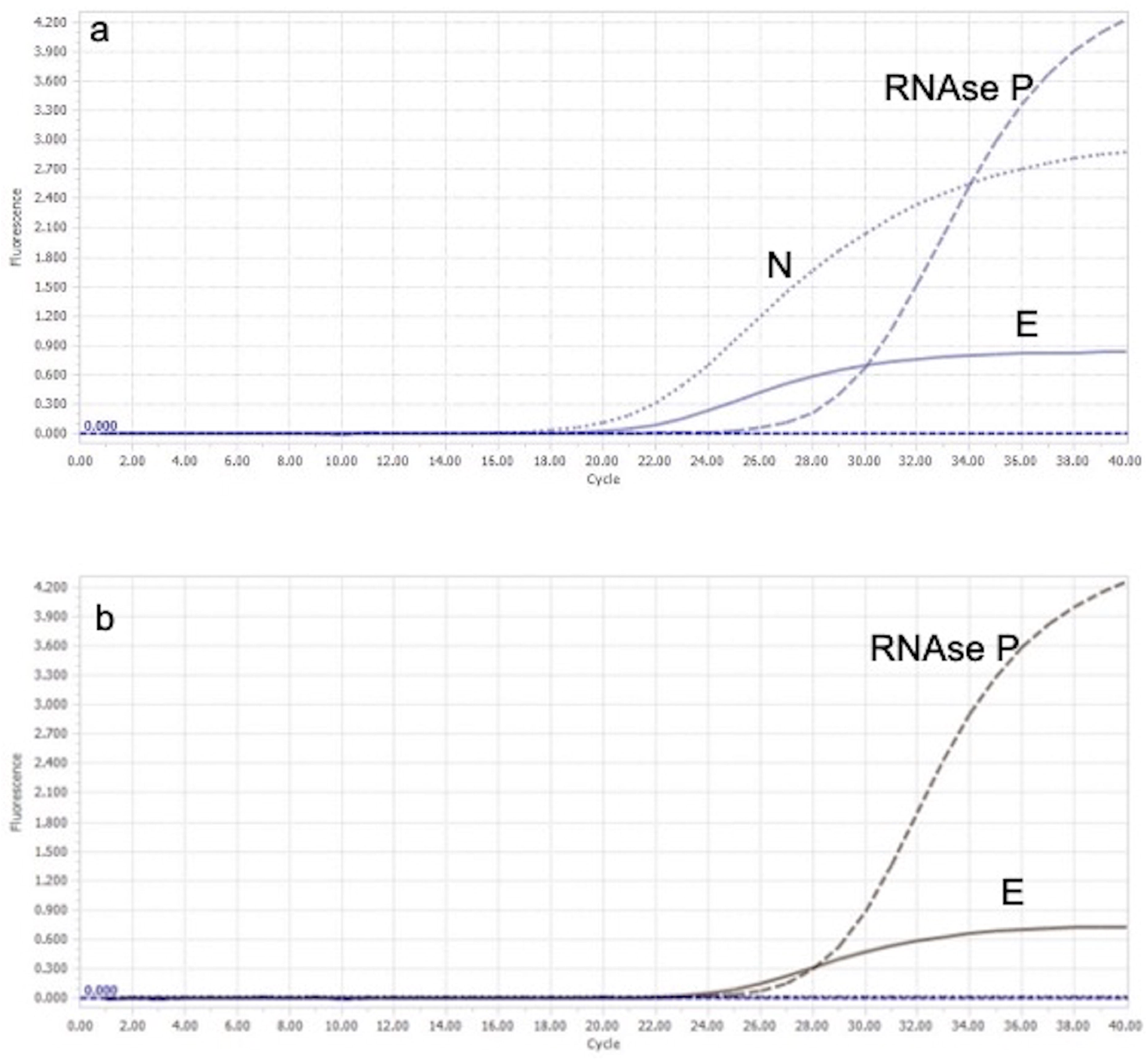
a: Case showing amplification in both E and N genes along with the internal control (RNAse P) amplification by RT-PCR. b: A case displaying N gene target failure (NGTF), with isolated amplification of E gene (analyzed on Light Cycler®96 Version 1.1.0.1320, Roche Diagnostics).

**Figure 3:**
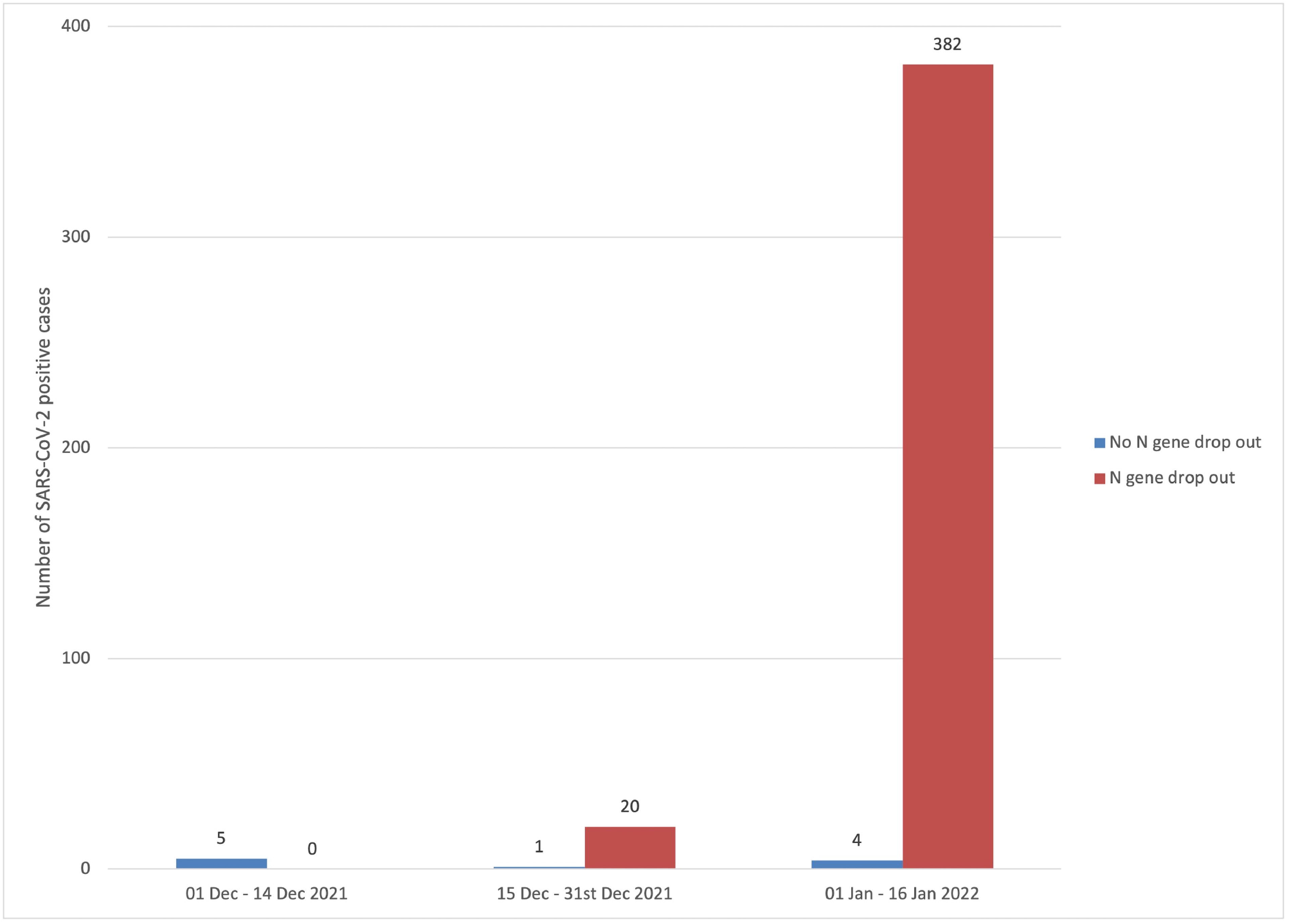
Comparative bar graph showing sharp rise in number of cases showing N gene drop out or N gene target failure (NGTF) compared to those without NGTF.

## 4. Discussion

The S gene target failure (SGTF) resulting from the HV69-70del is predominantly considered for identifying Omicron variant by most commercial kits [9]. However, mutations also affect other genes including ORF1ab, E and N in the Omicron variant [3]. Our findings confirm that the cases showing N gene dropout on RT-PCR (Huwel Life Sciences) are consistent with the Omicron variant, BA.1 lineage. The N gene probe overlaps with the ERS31-33del observed in Omicron, thus explaining the N gene target failure phenomenon (Figure 4). An important point here is that the S gene HV69-70del, responsible for SGTF, is not specific for the Omicron variant and is also observed in the Alpha (B.1.1.7) and Eta (B.1.525) variants, whereas the N gene ERS31-33del resulting in NTGF described here is not seen in any other SARS-CoV-2 variants of concern or interest. These findings reflect that Omicron variant can be reliably reported in cases showing NGTF by the current kit (Huwel Life Sciences), while dual amplification in E gene and N gene (or rarely N gene alone) is likely to represent the presence of delta/ non-Omicron variant. This separation does hold a clinical value in view of the reported differences in clinical severity and communicability between the two variants [10].

**Figure 4:**
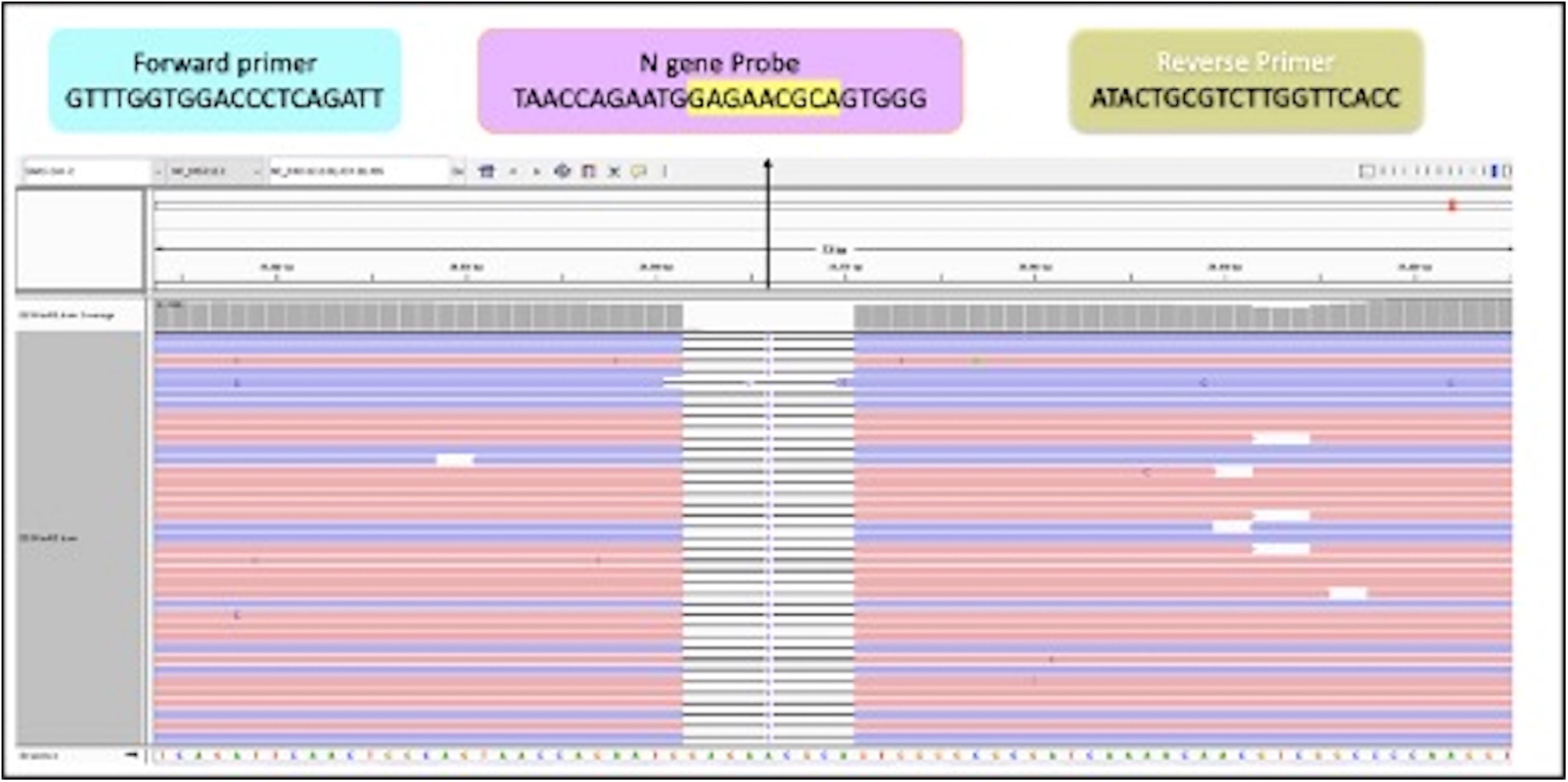
Aligned sequence reads in a case showing N gene drop out, depicting the presence of 9 nucleotide deletion (NC_045512.2:28,362-28,370) as seen in Integrated Genome Viewer (IGV) tool for visualization of indexed BAM files. Alignment was done against alignment with SARS-CoV2 reference genome (NC_045512.2). The top panel shows the N gene primer and probe sequences used in the commercial RT-PCR kit (Huwel Life Sciences). The presence of this deletion overlaps with the highlighted sequence N gene probe.

The second Omicron variant of the BA.2 lineage, termed ‘stealth’ Omicron, is genetically distinct from the BA.1 lineage and reported to escape detection on routinely used PCR kits as it does not harbour HV69-70del, therefore not resulting in S gene dropout [6] [11]. We did not encounter Omicron BA.2 lineage in our sequenced cases, however, we are confident that the NTGF by our RT-PCR method can be utilized to detect the BA.2 lineage as well, as the ERS31-33del in N gene is observed in both BA.1 and BA.2 lineages of Omicron variant [3]. We also emphasize that preservation of N gene or its dropout would depend on further viral genetic mutations and the probe design used for the gene in individual kits and that these findings concern the NTGF in the present kit used in the current clinical scenario only.

Our findings strongly highlight the possible fluctuations in the interpretation of RT-PCR results which can be encountered by laboratories involved in SARS-CoV-2 detection worldwide, especially as new SARS-CoV-2 variants with higher mutation load emerge in future. Following proper validation, the channelization of available commercial RT-PCR kits can be used as surrogates for detection of Omicron variant or any future variant at community-scale, thus reducing the burden on sequencing laboratories.

## 5. Conclusion

We describe a readily available RT-PCR strategy for rapid diagnosis of Omicron variant based on N-gene amplification failure.

The authors declare that they have no competing financial and non-financial interests.

## Data Availability

All data produced in the present study are available upon reasonable request to the authors

